# Large-Scale Measurement of Aggregate Human Colocation Patterns for Epidemiological Modeling

**DOI:** 10.1101/2020.12.16.20248272

**Authors:** Shankar Iyer, Brian Karrer, Daniel Citron, Farshad Kooti, Paige Maas, Zeyu Wang, Eugenia Giraudy, Ahmed Medhat, P. Alex Dow, Alex Pompe

## Abstract

To understand and model public health emergencies, epidemiologists need data that describes how humans are moving and interacting across physical space. Such data has traditionally been difficult for researchers to obtain with the temporal resolution and geographic breadth that is needed to study, for example, a global pandemic. This paper describes Colocation Maps, which are spatial network datasets that have been developed within Facebook’s Data For Good program. These Maps estimate how often people from different regions are colocated: in particular, for a pair of geographic regions x and y, these Maps estimate the probability that a randomly chosen person from x and a randomly chosen person from y are simultaneously located in the same place during a randomly chosen minute in a given week. These datasets are well suited to parametrize metapopulation models of disease spread or to measure temporal changes in interactions between people from different regions; indeed, they have already been used for both of these purposes during the COVID-19 pandemic. In this paper, we show how Colocation Maps differ from existing data sources, describe how the datasets are built, provide examples of their use in compartmental modeling, and summarize ideas for further development of these and related datasets. We also conduct the first large-scale analysis of human colocation patterns across the world. Among the findings of this study, we observe that a pair of regions can exhibit high colocation despite few people moving between them. We also find that although few pairs of people are colocated for many days over the course of a week, these pairs can contribute significant fractions of the total colocation time within a region or between pairs of regions.

## 1. Introduction

The worldwide use of mobile phones generates rich data describing human mobility, and epidemiologists have emphasized that this data can be vitally important for understanding the spread of infectious disease [1, 2, 3]. During the ongoing COVID-19 pandemic, such data has been used to parametrize compartmental models [4, 5] and to measure the causal impact of mobility-oriented interventions [6, 7, 8, 9, 10, 11]. However, this data is usually only available to researchers for specific parts of the world at specific times, through limited agreements with regional mobile-phone providers. In a localized epidemic, there may be no mobile-phone provider that is willing or able to provide such data, and in a global pandemic, there may be no way to obtain data with worldwide breadth.

Over the past three years, the Facebook Data for Good program has made aggregated mobility datasets, based upon the mobility traces of consenting Facebook users and with privacy protections applied, available to humanitarian organizations who are responding to natural disasters around the world [12]. In this paper, we describe Colocation Maps, our first map built in consultation with epidemiologists and tailored specifically to the epidemiological use case. Colocation Maps answer the following question: for a pair of geographic regions x and y (e.g., counties in the US), what is the probability that a randomly chosen person from x and a randomly chosen person from y are simultaneously located in the same small region (approximately 0.6km × 0.6km squares) during a randomly chosen minute in a given week? Figure 1 shows an example colocation network for Italy, where the links between regions are weighted by the probability defined above.

**Figure 1.**
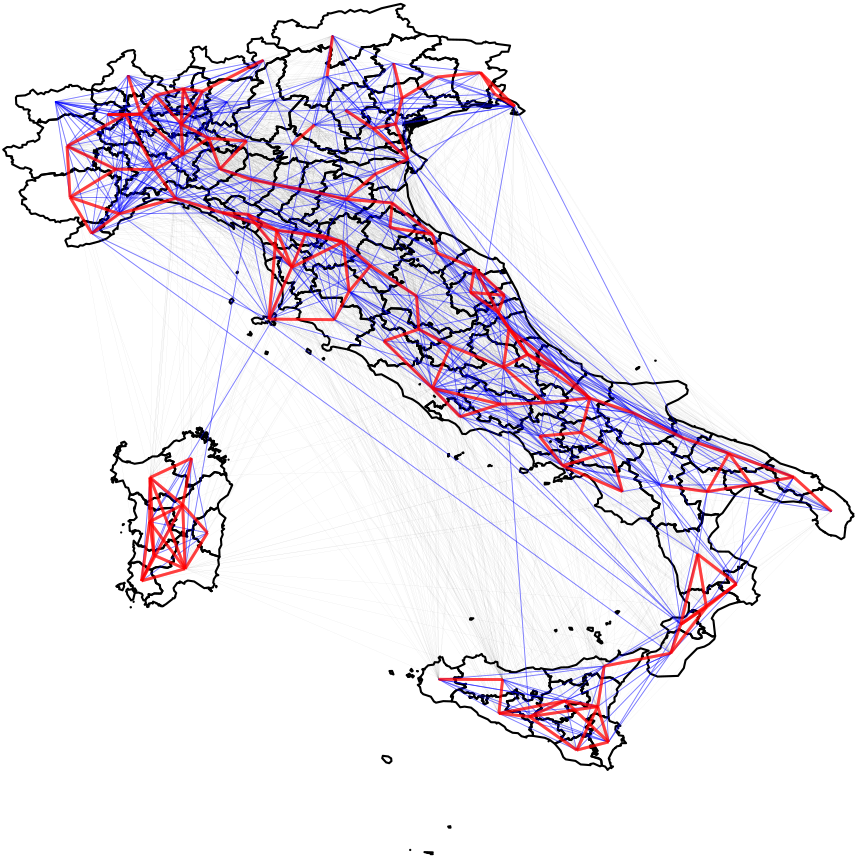
Example Colocation Map for Italy for the week of 2020-02-26 to 2020-03-03. Red links correspond to the strongest colocation ties, blue links indicate intermediate ties, and black ties are the weakest.

Estimating colocation is especially useful for epidemiological modeling of human-to-human disease transmission. Such modeling requires an understanding of *mixing patterns* (i.e. how often people could transmit a disease by coming into contact with other people). Colocation Maps can provide such an understanding, and directly inform *metapopulation models* of disease spread. In these models, epidemiologists consider various subpopulations representing different geographic regions, and the level of coupling between subpopulations depends on how often people from each region come into contact that is sufficient to transmit a disease. Epidemiologists often indirectly parametrize these couplings through data that do not actually represent contact rates, such as counts of people who move between locations (e.g., travel or commuting data) [13, 14, 15]. Colocation Maps can offer a better parametrization and one that can evolve temporally over, for example, the course of the COVID-19 pandemic, where policy and mobility patterns will continue to evolve rapidly.

While Colocation Maps were first shared with epidemiologists in April 2020 [16], this is the first paper to describe them in detail and demonstrate how they can be used to model the spread of contagious disease. This paper is intended both as a resource for epidemiologists and other researchers who use Colocation Maps and as an exploration of human colocation patterns. We begin, in the Materials and Methods Section, by describing the underlying data source and specifying the transformations performed upon that raw data to produce Colocation Maps. We continue in the Applications of Colocation Maps section by providing a derivation of a simple metapopulation SIR model parametrized with Colocation Maps (thus demonstrating how the datasets can be applied in practice), by showing how colocation data differs from movement data (thus showing how these datasets can yield different insights than traditional mobility datasets), and by surveying work from research groups that leveraged Colocation Maps during the early months of the COVID-19 pandemic. A key finding in this section is that pairs of regions can exhibit high colocation despite there not being much direct movement between the regions. We continue in the Assumptions of Colocation Maps section by reporting analyses that test the underlying assumptions of Colocation Maps. Along the way, we find that a small fraction of pairs of people that are frequently colocated can nevertheless account for a large fraction of total colocation time. In our concluding section, we propose directions for further development of these datasets.

## 2. Materials and Methods

### 2.1 Data Sources

#### Facebook Location History Data

Generally speaking, the term “human mobility data” means any data about how people move through physical space [17]. For the purpose of this paper, mobility data specifically refers to location updates from a mobile phone in the form of longitude *x* and latitude *y* coordinates at a given time *t*, where such an update could be written (*x, y, t*). Facebook collects mobility data from people who have explicitly chosen to enable location services for Facebook applications. In particular, Colocation Maps utilizes mobility data from the main Facebook application on iOS and Android, and only includes data from people who opt in to Location History (LH) and Background Location collection (BC)^1^.

People specify their Location Settings in the app and can change them at any time. In July 2020, for those with LH and BC enabled, 54% of the population provided a location update in at least half of the five-minute intervals covering each day. This coverage varied by time of day, with daytime having 58% of people with location updates in at least half of five minute intervals; while at night only 46% did. Use of this pair of settings varied by country, with adoption at a median of 5.2%, ranging from less than 1% to 8.4% for the 10th and 90th percentile countries, respectively.

#### Comparison of Location History Data to Other Data Sources

Traditional data sources for human mobility data include censuses [18, 19, 20], local surveys [21, 22], and travel demand data from airlines or other transportation providers [23, 24, 25, 26]. In recent years, popular data sources include call detail records (CDR) from mobile phone providers [21, 20, 27, 28] and global positioning system (GPS) traces [29]. Data from location-based services on web platforms (e.g., Twitter) has also been used [18, 30, 31, 12].

Facebook LH data has spatial and temporal resolution that is comparable to GPS data, but with greater international breadth. The spatial resolution is finer than many CDR datasets, which aggregate up to the cell-site level. Also, importantly, Colocation Maps transform the raw LH data in a manner that is more aligned with the needs of metapopulation modeling than GPS or CDR datasets, which are often shared as matrices of transition counts between locations. This type of aggregation does not necessarily approximate the amount of contacts that could result in disease transmission. Colocation Maps instead aggregate the raw data (on Facebook’s servers) so as to bridge the gap between these transition-count datasets and datasets that directly measure face-to-face contact. Except in certain limited contexts where agreements are made with small populations [32], datasets describing very close-range face-to-face interactions are difficult and invasive to obtain. By measuring when people are colocated within a somewhat larger spatial scale, Colocation Maps can provide data that more closely approximate disease-transmitting contacts than transition matrices and do it at a scale that would not be possible when measuring actual face-to-face contact.

Colocation Maps are built from data from online social network users, and researchers have expressed concern about how well such data represents the population on the ground [17]. Researchers have indeed found that Twitter users who share their location constitute a biased slice of the population [33]. We explore the representativeness of Colocation Maps datasets further in the Assumptions of Colocation Maps section. However, it is worth noting that representativeness questions appear in different forms with all of the traditional sources of human mobility data listed above. Surprisingly and perhaps reassuringly, researchers have found that mobility inferences gleaned from different sources are often consistent [18].

#### Administrative Polygons

Epidemiological models can incorporate various levels of detail in the contact structure between individuals, ranging from whole-population models (where everyone in the population is assumed to interact with everyone else at the same rate [34]) to very detailed agent-based models [35, 36]. Colocation Maps are intended to be most directly applicable to metapopulation models, which sit between these extremes. In these models, the full population of interest is divided up into subpopulations. The rate at which a person in subpopulation *i* interacts with someone in subpopulation *j* can be different for each pair of subpopulations *i j* [37]. Often, the subpopulations correspond to geographic regions, and in these circumstances, it is convenient to use regions that correspond to political administrative units, because these are the geographic units to which interventions can be most easily delivered. Generally, public health decisions (or relevant data, such as disease incidence numbers) occur within countries, at the county-scale. Therefore, Colocation Maps are aggregated at the level of administrative regions on the scale of counties or cities. In particular, our administrative boundaries are Pitney-Bowes polygons (a commercially available set of polygons) at level three if possible, otherwise level four [38]. In the United States, level 3 divides the country into 3,142 counties, and at level 4, the country is represented by 35,502 towns. Administrative regions at a larger scale, such as states or countries, would be a less accurate description of mixing patterns and less actionable. Meanwhile, if we were to aggregate at a finer scale, we would need to discard more data to remain in accordance with the privacy principles that we describe below.

### 2.2 Dataset Preparation

In this section, we describe the series of steps that lead from raw LH data to Colocation Maps.

#### Defining the Relevant Area

To build a Colocation Map, we first need to specify an area of the world to study. This area is determined by the specific modeling goals of the research team that is requesting the data. When we receive a request for data, we leverage the existing infrastructure from the Disaster Maps component of our Data for Good program, where we draw a rectangular bounding box that encapsulates the area affected by a natural disaster. In the Disaster Maps context, this bounding box defines a Facebook Data For Good *crisis* [12]. In the Disease Prevention Maps context, the term “crisis” is not always relevant (e.g., if epidemiologists use the data to study hypothetical scenarios for preventative research), so we use the term “project” instead to refer to the instantiation of the bounding box in our pipelines. The Pitney-Bowes administrative polygons are then intersected with the bounding box for the project, and any geographic region that overlaps with the bounding box is included in the resulting map. Example bounding boxes enclose a geographical region containing an entire country, the data from which could serve to support epidemiological research on a country scale.

Once a project area is specified through the creation of a bounding box, Colocation Maps are automatically produced on a weekly basis for that region, where each week’s computation is entirely independent, except for utilizing the same bounding box. When we refer to a singular map, we mean a single week’s computation for a particular project.

#### Identifying a Home Population for an Administrative Region

Our first task in computing colocation is to assign home regions to those people who have LH and BC enabled. Even among this population, mobility data may be incomplete, with gaps between location updates. This can occur for a variety of reasons, such as turning off one’s phone, a lack of cell phone coverage, and differences in behavior between iOS and Android. Thus, we only include people in the calculation if their mobility trace over the calculation satisfies certain properties, which we will motivate as we describe the calculation below.

Home-estimation approaches typically focus on people’s mobility data at night, since it is expected that people are likely to spend the night near their homes [39]. We define the “night” of a specific date (e.g., 2020-03-03) to correspond to 20:00 (8pm) on the previous calendar day (e.g., 2020-03-02) to 06:00 (6am) on the calendar day itself (see panel a of of Supplementary Figure S1 for an illustration of our division of dates into day and night). We map pings to local times (and subsequently classify them as daytime pings, nighttime pings, or neither) based on the geographic location where the ping was recorded; for instance, if a ping was recorded at 9pm in New York, that would be considered a nighttime ping, but a ping that was recorded at the same time in California would *not* be considered a nighttime ping, since it would only be 6pm there. If a person logged at least three nighttime pings for a given date, we compute a nightly modal region for that person for that date. We consider all of an individual’s nightly modal locations over a 10-date interval around the date of the computation and assign a home region to the individual if the same region is their nightly modal region on at least 6 nights. This process is illustrated in panel b of Supplementary Figure S1. If an individual does not have a consistent nightly modal location over six nights, then they are not included in the calculation.

#### Trajectory Construction

##### Who has sufficiently complete trajectories?

Our next task is to construct trajectories for the remaining people in the computation. Eventually, we will need to intersect trajectories to determine if pairs of people were colocated. However, as noted above, we may only have irregular and intermittent location updates for any given person, so it is not possible to assert whether two people were colocated at **precisely** the same time. To resolve this, we divide the week into five minute bins, and our definition of colocation is that two people are within the same level 16 Bing tile (approximately 600 meter by 600 meter squares, at the equator) within the same five minute bin. Even with this binning, location updates for an individual may be too sparse in time to credibly reconstruct a trajectory. Therefore, we place conditions on the observed frequency of location updates over the period of the calculation. Because the collection rate of LH pings differs between the day and night, we introduce different conditions for different parts of the day. In particular, we require that an individual have a location update in at least 9 of 10 hours during the day, 4 of 10 hours at night, and 17 out of 24 hours over the full 24 hours corresponding to a date. We further require that the individual satisfy these three criteria on all 10 days of a period including the calculation date. See panel c of Supplementary Figure S1 for an example. People who do not satisfy these criteria are excluded from the calculation.

For the remaining individuals, we construct observed trajectories, consisting of temporal sequences of (x, y, t) points, from the LH data. Although we have focused on *local* days and *local* nights up to this step in the calculation, we now shift to constructing trajectories for a fixed one-week window, spanning the seven days up to and including the calculation date in Pacific time. There are three reasons for this:

1. Averaging over any contiguous week averages over the same number of daytimes, nighttimes, etc. everywhere in the world.
2. Bracketing trajectories for individuals by the local time of pings can introduce anomalies that can lead to inconsistencies in the calculation (e.g., if we end the calculation at midnight on a given date, but then an individual travels across a time zone barrier between 12pm and 1am, they can have another ping that falls within the allowed local time window).
3. It is much more computationally efficient to work with a universal, fixed-time window.

##### Speed-based Filtering of Trajectories

We next filter trajectories that include unrealistically fast movement (e.g., where the person seems to move at thousands of miles an hour multiple times). We have observed examples of this in the data, where many people seem to “teleport” to a single level-16 Bing tile many times. This can be a data quality concern for Colocation Maps: in particular, it would result in a large number of (likely) inaccurate colocation events. To combat this, between every consecutive pair of observed location updates (*x*_1_, *y*_1_, *t*_1_) and (*x*_2_, *y*_2_, *t*_2_) in a trajectory, we estimate the average speed required to go from one update to the other and record that transition as a speed outlier if it is above 200 MPH. We illustrate an example of this in panel d of Supplementary Figure S1. We exclude any trajectory that has five or more speed outliers over the week.

##### Trajectory imputation

After filtering trajectories based on update frequency and speed, we complete the remaining trajectories. Because we have imposed requirements on the frequency of pings in a time interval that includes the seven-day colocation period, we are guaranteed that trajectories are reasonably complete for the people remaining in our computation. Nevertheless, there can still be many missing five-minute bins for which we need to impute locations. We utilize a simple imputation strategy that asserts that an individual remains stationary until halfway between observed location updates. To be specific, if an individual has two consecutive location updates (*x*_1_, *y*_1_, *t*_1_) and (*x*_2_, *y*_2_, *t*_2_) where *t*_2_ > *t*_1_ and there are *n*_21_ five-minute bins between *t*_1_ and *t*_2_, we impute that their location is (*x*_1_, *y*_1_) for the first 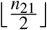 of the intervening five-minute bins and that their location is (*x*_2_, *y*_2_) for the last 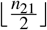 of the intervening five-minute bins. If *n*_21_ is odd, then there will be one five-minute bin that is equidistant in time from the observed bins, and we break that tie randomly to impute one of the two observed locations. This process is illustrated in panel d of Supplementary Figure S1.

#### Estimating Colocation Probabilities

We now have complete week-long trajectories for all people who remain in the calculation, and we can therefore compute *X*_*i jr*_, the number of people assigned to administrative region *r* who are located within level 16 Bing tile *i* during five minute time bin *j*. From this, we can in turn calculate *m*_*rs*_, the number of weekly colocations between two regions *r* and *s*:

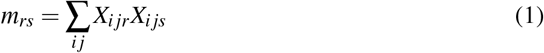

The sum over *i* iterates over all possible level 16 Bing tiles, and the sum over *j* iterates over all 2016 five-minute time bins in the week under consideration If *r* = *s*, we instead compute *m*_*rr*_ = ∑_*i j*_ *X*_*i jr*_(*X*_*i jr*_ −1) to avoid counting a user as colocated with themselves. Following network-science convention for adjacency matrices, this intentionally double counts the number of colocations for within-region colocations [40]. This process of counting the number of people in each Bing tile in each time bin and then performing the sum in equation (1) is the computationally tractable way of performing, at scale, the trajectory intersections illustrated in panel e of Supplementary Figure S1.

The result of a Colocation Map calculation is a probability matrix, where matrix element *p*_*rs*_ is the probability of colocation between region *r* and *s*. We compute this probability from *m* as follows

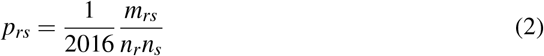

for distinct regions *r* and *s*. If *r* = *s*, we divide by *n*_*r*_(*n*_*r*_ −1) instead of *n*_*r*_*n*_*s*_. Regardless of whether *r* = *s* or not, this probability can be interpreted as the number of realized colocation events in the numerator divided by the number of possible colocation events in the denominator. It hence is correctly normalized to the range of zero to one.

The matrix *p*_*rs*_ is shared with partners through a secured portal called GeoInsights, which we will describe below. Partners can inspect the matrix within GeoInsights (Supplementary Figure S2 shows an example visualization in the tool), or download a CSV file that contains one row per pair of polygons *r* and *s*. Supplementary Table S1 enumerates the columns that are included in the CSV and their meanings.

#### Privacy Protections

We will now discuss the principles around how we think about privacy in the context of Colocation Maps and, in particular, how the quantity *p*_*rs*_ is defined with privacy considerations in mind. Privacy researchers have consistently found that *individual-level* human-mobility data is vulnerable to reidentification attacks, even when all explicitly identifiable information (e.g., names, social security numbers, etc.) is removed. Here, a reidentification attack refers to a situation where an attacker knows some mobility information about an individual, uses that information to identify that person’s records in a dataset, and thereby learns new information about that target individual [41]. Because mobility traces are highly unique across individuals, this type of reidentification is often possible in individual-level datasets. For example, in 2011, Zang and Bolot published a study of three months of CDRs for 25 million mobile-phone users in the United States, finding that 50% of people are unique once their top three most frequently visited locations are specified at the cell-site level. Meanwhile, de Montjoye et al. also looked at CDR data at the cell-site level, but considered trajectory data, where the dataset reports precise times at which an individual visited a particular location. These authors found that 95% of people can be uniquely identified by four spatiotemporal points [42].

Due to the vulnerabilities of individual-level mobility data, within Facebook, this data is stored in anonymized form, is accessible only to developers who are working on specific products, and is permanently deleted within 60 days. Meanwhile, the datasets that we share with humanitarians and epidemiologists through the Data for Good program are all de-identified and aggregated over many individuals [12]. However, it is important to note that, with certain aggregation approaches, substantial reidentification risk can remain after aggregation.This is due to three empirically well-established properties of human mobility:

1. human mobility is highly diverse across individuals
2. human mobility is highly recurrent, day over day, for the same individual
3. people tend to demonstrate little mobility at night, and their mobility during the day is continuous (i.e., people cannot teleport)

Xu et al. have shown that these three properties can be leveraged to reconstruct accurate trajectories from datasets that report the aggregate number of people observed in a given location at a given time [39]. These reconstructed trajectories are then vulnerable to reidentification attacks, along the lines of [43] and [42].

Colocation Maps are designed with this vulnerability in mind. Recall that *p*_*rs*_ estimates the amount of time a randomly chosen person from region *r* spends near a randomly chosen person from region *s* over the course of a week. By reporting this rate at a temporal granularity of a week, Colocation Maps average over the diurnal rhythms in people’s movements, and it is precisely these rhythms that make human-mobility data such distinctive digital fingerprints of individuals. Furthermore, the maps do not indicate where or precisely when colocation events occur: the colocation rate for Los Angeles County and San Francisco County sums over colocation events that happen in Los Angeles, in San Francisco, and in all other administrative polygons (e.g., if an individual from Los Angeles and an individual from San Francisco happen to be near one another in Yosemite or in New York). Finally, as in our other Disaster and Disease Prevention Maps [12], we aggregate up to polygons that are generally on the scale of counties in the United States and drop data for polygons that are underrepresented in our data (i.e., that are the home polygons of fewer than 10 people). This means that the end user of the dataset can tell that there were *n*_*r*_ people whose (inferred) home location was Los Angeles County (where *n*_*r*_ ≥10) but cannot even identify, with complete certainty, a second location that any of those people visited over the course of the week.

#### Facebook’s Data for Good Program

The privacy protections that we have built into the construction of Colocation Maps are meant to mitigate the risk that an individual can be explicitly reidentified in the data. However, as has been noted in the literature, statistical privacy protections are not meant to prevent the end user from learning general behavioral trends for the population [44, 45]. Indeed, learning these trends is the very point of dataset release. Thus, to ensure that these trends are only learned by trustworthy end users, procedural privacy protections are necessary as well. Such protections are managed through Facebook’s Data for Good program [46].

The Data for Good program makes data sets available taking direction from general guidelines provided by civil society and academic data privacy advocates [47]. Public health experts [2] were consulted extensively to define the specific colocation data likely to provide efficacy for pandemic response. Access to Colocation Maps is available to vetted non-profit organizations and academic research institutions through Facebook’s Data for Good program. Partners only have access to de-identified and aggregate information from Facebook and thus no individual level information is shared. Data sets, including Colocation Maps, are made available through the Data for Good program under the terms of a data license agreement which defines the allowed terms of use by partners.

Once a partner institution’s request for access is vetted and an appropriate data license agreement is signed, then access is granted through a Facebook’s web-based spatial visualization tool called GeoInsights. An example visualization of colocation data in GeoInsights is shown above in Figure 2. GeoInsights also allows data to be downloaded by partners in a variety of file formats^2^.

**Figure 2.**
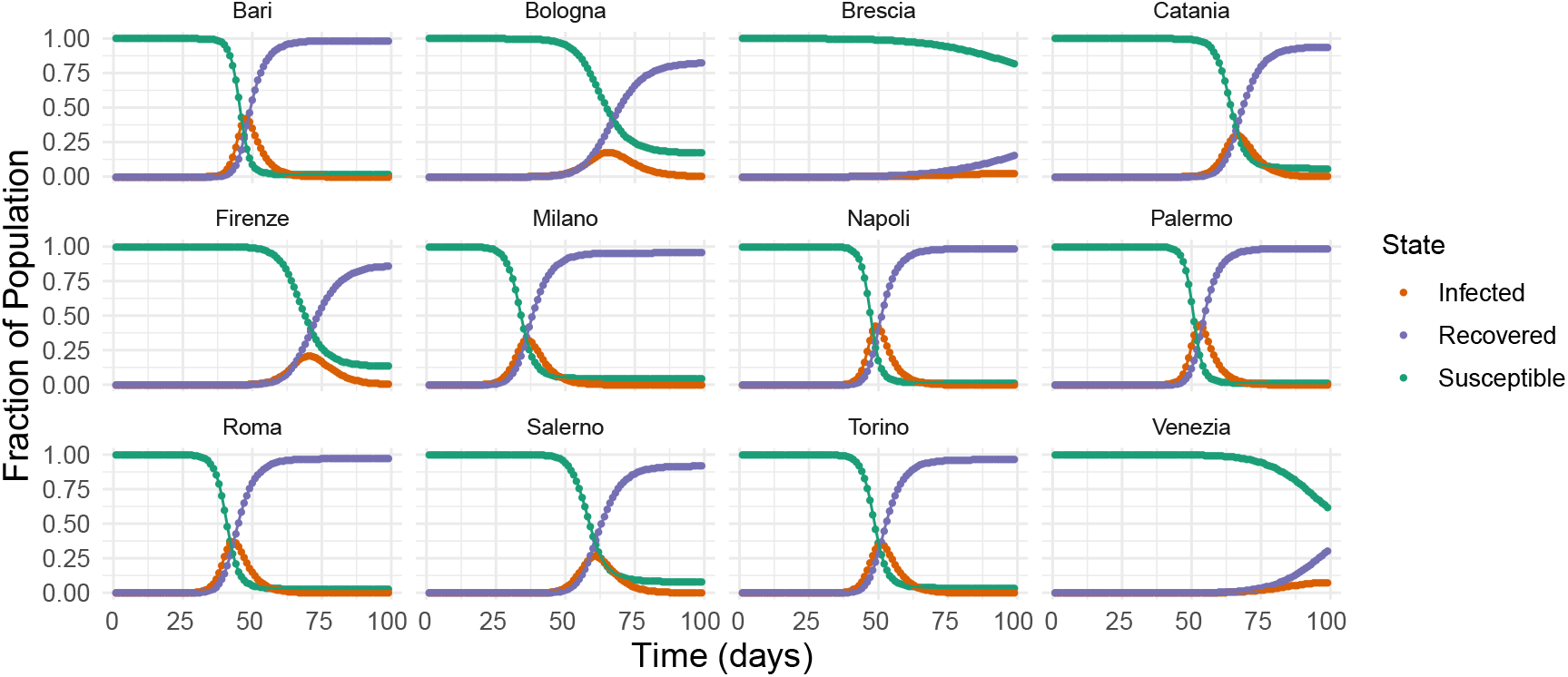
Example trajectories from Eq. 6, adapted to represent metapopulation SIR infectious disease dynamics for the provinces of Italy. The initial outbreak occurs in Milan, and we plot the population fractions of susceptible, infected, and recovered individuals over the course of 14 weeks for 12 provinces.

## 3. Applications of Colocation Maps

We now demonstrate why Colocation Maps are useful for epidemiological modeling. First, we provide an example of how these datasets can be used to parametrize compartmental models. Next, we empirically demonstrate that the mixing patterns captured by Colocation Maps are not well represented by measurements of movement between regions, and therefore, studies that leverage Colocation Maps can lead to different insights than those that utilize conventional human-mobility data sources. Finally, we survey work that has leveraged Colocation Maps in the first months of the COVID-19 pandemic.

### 3.1 An Example Metapopulation Model

We now provide a derivation of a simple *SIR* metapopulation model using the probabilities provided by Colocation Maps from a network model of disease spread. We do not imply here that this model accurately predicts the spread of any disease, and instead, we include this here purely to give an example that researchers can reference when parameterizing more practically relevant models.

We assume we have an individual-level colocation network *G* with time-varying adjacency matrix given by *A*_*i jk*_ where *A*_*i jk*_ = 1 if user *i* and *j* are colocated during five-minute time bin *k* and zero otherwise. Let *P*(*S*_*i*_), *P*(*I*_*i*_), *P*(*R*_*i*_) be the time-varying marginal probabilities that vertex *i* is susceptible, infected, and recovered in an SIR model, where *β* is the constant rate at which infection is spread from an infected to susceptible individual while two individuals are colocated, and *γ* is the constant rate at which a person recovers. Ignoring correlations (i.e., utilizing the naive mean-field approximation, in which we assume for example that *P*(*S*_*i*_, *I*_*j*_) ≈ *P*(*S*_*i*_)≈*P*(*I*_*j*_)), we have the following nearly standard set of differential equations:

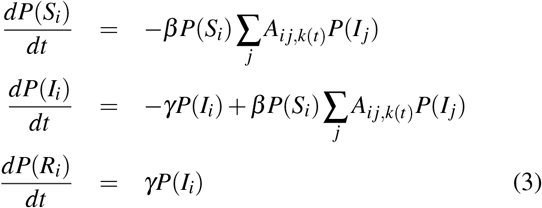

where *k*(*t*) is the five minute time bin associated with time *t*. We cannot actually utilize this model because we do not know *A*_*i j,k*(*t*)_ for the population. To arrive at a usable model, we assume each user has been assigned to a geographic region, and make the following homogeneity approximation for a user *i* who has been assigned to region *r*

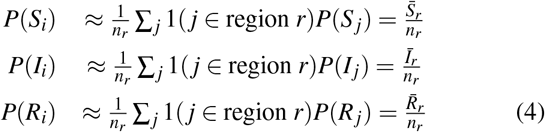

where the second equality defines 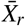 as the expected number of users assigned to region *r* in state *X*.

Writing the dynamical equation for 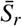 using this assumption, we find:

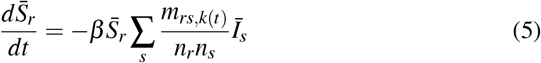

where *m*_*rs,k*(*t*)_ is the number of colocations between region *r* and *s* (twice this if *r* = *s*) during time bin *k*. Details of this calculation can be found in the Supplementary Information. We now make an additional approximation that *m*_*rs,k*_ ≈ *p*_*rs*_*n*_*r*_*n*_*s*_^3^. Essentially, this asserts that the expected number of colocations between two regions is uniform across time bins during the week. Using this additional approximation yields:

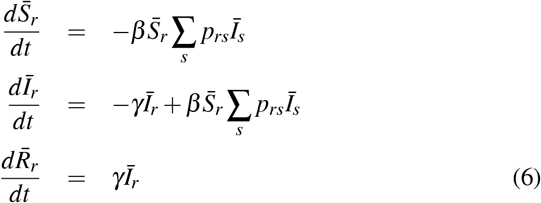

This metapopulation SIR model is defined entirely in terms of the probabilities provided by Colocation Maps and the *SIR* parameters *β* and *γ*.

In Figure S3, we plot example trajectories from an SIR metapopulation model, realized by adapting equations (6) to predict the spread of an infectious disease through Italy. Each sub-population represents one of Italy’s 110 provinces. For this example, we choose *β* = 4 × 10^−4^ and *γ* = 0.25, and initialize the simulated outbreak to start with 10 individuals in the province of Milano. We simulate the disease spread starting in the week of March 3, 2020 through the week of June 6, 2020, using each week’s colocation matrix to represent the interaction strengths between individuals in different provinces. In Supplementary Figure S3, we also plot the first time at which the number of infected individuals reaches 100 in each province, plotted against the log of the colocation strength between that province and Milano. We observe that the simulated epidemic more quickly reaches provinces which interact more strongly with Milano.

### 3.2 Colocation vs. Movement

The datasets that are often used to parametrize compartmental models such as (6) measure the number of people who are moving between pairs of places. How is the colocation rate between a pair of places related to the movement flux between those places? If colocation is highly correlated with movement flux, then perhaps Colocation Maps would not add much information beyond traditionally available datasets. However, this turns out to not be the case; there can be large variance in the level of colocation over pairs of regions that have similar amounts of movement flux, especially when the movement flux is low.

To demonstrate this, for several regions of the world, we construct a weighted, undirected “colocation network” and a weighted, unirected “movement network” for the week ending 2020-08-11. In both networks, the nodes represent administrative regions but the edges between these regions are weighted differently. In the colocation network, each edge is weighted by *m*_*rs*_, the number of colocation events between people from the regions during the week. In the movement network, each edge is weighted based on data from Facebook Movement Maps, which measure, in eight-hour intervals, the number of people who move between regions [12]. The edges of the movement network are weighted by the maximum movement flow between the two regions (in either direction) over the week.

Figure 3 shows distributions of the log of the number of colocation events, cut by the max movement flow, for 18 different colocation maps. In the legend, within parentheses, we also include the Kendall *τ*_*b*_ correlation between the movement and colocation weights within each country. This plot shows that:

**Figure 3.**
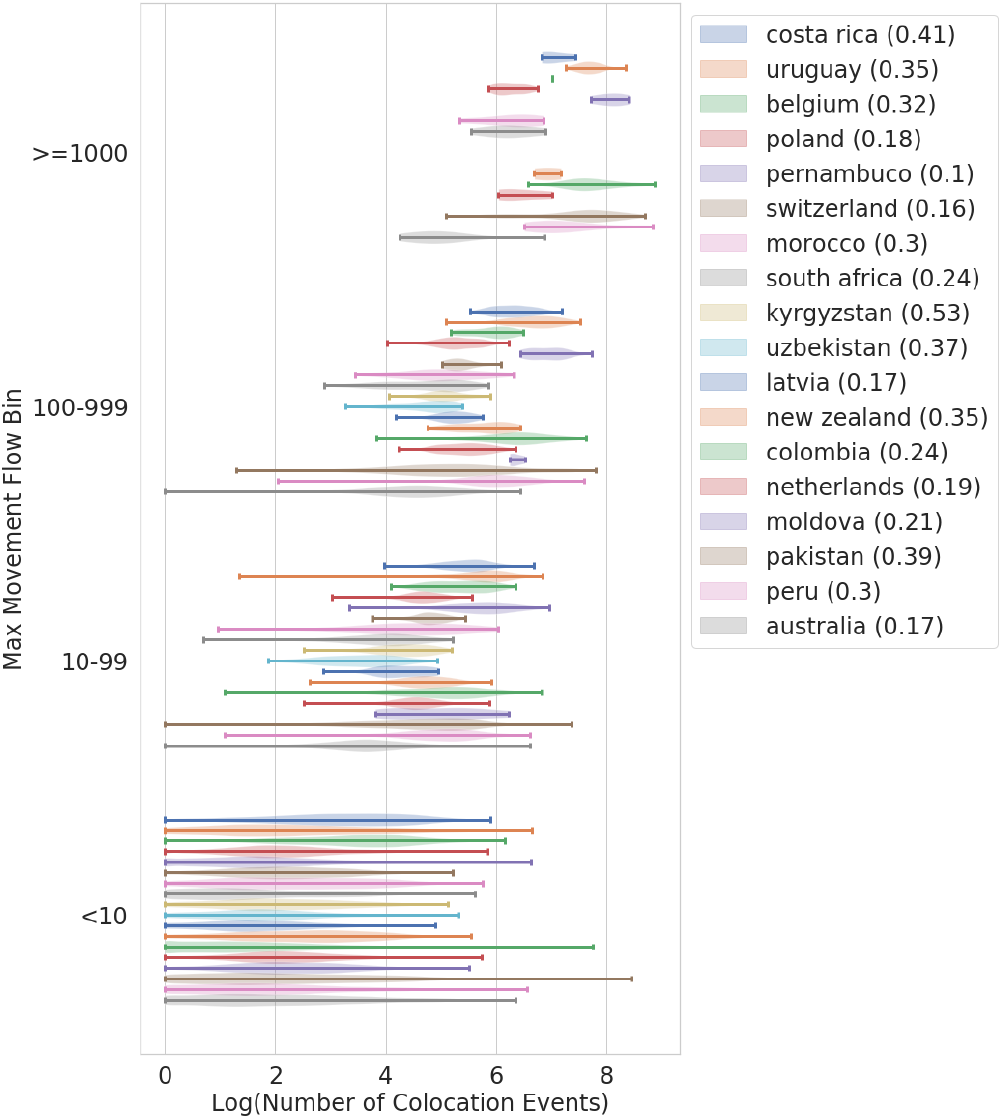
Distributions of the number of weekly colocation events over polygon-polygon pairs between which there is a given level of movement. For example, the 10-99 bin along the y-axis includes all polygon-polygon pairs such that the maximum observed movement between the polygons was between 10-99 people. Movement here refers to the number of people who were observed in the first polygon during an 8-hour bin during a given week and then in the second polygon during the next 8-hour bin. Each violin in the 10-99 bin shows the distribution, in a given region of the world, of the log of the number of colocation events between the polygons in the pair. A colocation event occurs whenever an individual from the first polygon and an individual from the second polygon are in the same level-16 Bing tile in the same five-minute window. Data is plotted for the week ending 2020-08-11 for 18 different regions of the world. For each region of the world, we also include in parentheses the Kendall *τ*_*b*_ correlation between movement and colocation weights within each country.

- Especially in the lowest movement bins, the distribution of the number of colocation events varies over many orders of magnitude, sometimes with non-negligible amount of weight of the distribution at relatively high values (e.g., *m*_*rs*_ ≥ 10^4^).
- The variance of the colocation distribution decreases as movement grows.
- The values of the Kendall *τ*_*b*_ correlation, while all statistically significantly different from 0, are nevertheless quantitatively weak, indicating that the movement values are not a good guide to even the rank ordering of the colocation values.

### 3.3 Applications of Colocation Maps to Research on COVID-19

Since Colocation Maps were made available to the research community in April 2020, they have been used by multiple research teams to understand the unfolding COVID-19 pandemic. Chang et al. used Facebook Colocation Maps and Movement Maps to model the spread of COVID-19 in Taiwan. They showed that, in terms of the total number of infections, reducing intra-city travel would have a stronger impact on spread than reducing inter-city travel, but that inter-city travel reductions are more effective in containing the scope of outbreak in one area and in helping officials target resources to the right location [4]. Meanwhile, researchers at Texas A&M conducted spatial network analysis of the temporal colocation network amongst United States counties and correlated trends in edge weights with trends in COVID-19 cases, finding that the colocation reduction is associated with a reduction in case growth with a week of delay [5]. Finally, multiple research teams have created dashboards and visualizations of the colocation data to track the spread of the disease:

- A team at the University of Adelaide used Colocation Maps to derive estimates for the average number of contacts per person in a given region with people in another region^4^.
- A team at Chapman University used colocation data to track the spread of COVID-19 in Hong Kong and to visualize the colocation probabilities among different regions^5^. The same team built another visualization showing the level of connectivity in different regions in Hong Kong to aid with COVID-19 response^6^.
- A team at the London School of Hygiene & Tropical Medicine used Colocation Maps to create a dashboard showing the colocation probability over time in the United Kingdom^7^.

## 4. Assumptions of Colocation Maps

Having demonstrated how Colocation Maps can be applied in practice, we now turn to analyses of the assumptions that are built into these datasets. These include the assumption of representativeness of our data, the assumption that colocation can approximate face-to-face contact, and the assumptions of spatial and temporal homogeneity that we used in our simple metapopulation model. These assumptions are, of course, not strictly true. However, the spatial and temporal homogeneity assumptions are not just features of Colocation Maps datasets, but are also built into most metapopulation models [37]. Indeed, they are what differentiate metapopulation models from more fine-grained modeling approaches, such as network modeling [48, 49] and agent-based modeling [50, 35]. Metapopulation models are useful because the additional refinement of these other modeling approaches is not always necessary for gaining meaningful understanding about the progression of an epidemic, and Colocation Maps datasets remain useful, despite the inexactness of their core assumptions, for the same reason. Moreover, we should also note up front that relaxing some of these assumptions, or defining colocation on finer spatial scales so as to get “closer” to face-to-face contact, entails tradeoffs with privacy. Despite these considerations, it is valuable to measure the extent to which the assumptions behind Colocation Maps hold, so that end users of the datasets are aware of the biases that may be present in the data and in order to formulate ideas for mitigating those biases.

### 4.1 Representativeness

As noted above, researchers have expressed concerns about the representativeness of datasets derived from usage of web products [18, 17]. In practice, all data sources for human mobility have their limitations. Censuses offer some of the most comprehensive surveys of the population and can even be used to glean insights about mobility (e.g., through commuting flows or year-over-year relocation counts), but they are infrequently conducted, not available for all countries, and cannot be used to pose questions like those addressed by Colocation Maps. By contrast, data from web products are inevitably biased towards that segment of the population that uses the internet most, but they can offer insights into how situations evolve on shorter time scales.

Our goal here is to give practitioners who are using Colocation Maps a window into *where* our data sets are likely to be most trustworthy. In terms of the number of users that are included in Colocation Maps, we have decent coverage for most countries in Europe, North America, South America, South Asia and Southeast Asia, but less coverage in other parts of Asia, Africa and Oceania. Figure 4 compares the age distribution of people who are included in Colocation Maps to distributions from the high-resolution settlement layer (HRSL) and from population data from the United Nations (UN)^8^ when HRSL is not available. The HRSL is a dataset, built by data scientists at Facebook based only on satellite data, which provides state-of-the-art estimates of population density in many regions of the world [51]. Due to the age restriction of Facebook users and the limit of HRSL, we only include population with age more than 14 years old. From Figure 4, the Colocation population is skewed towards older population (defined as age more than 29 years old) in most regions of the world, which might be contrary to the general impression that Internet is for young people. Supplementary Figure S4 performs a similar comparison for gender and shows that the Colocation population is skewed towards female in South America and East Europe, and skewed towards male in South Asia and Africa.

**Figure 4.**
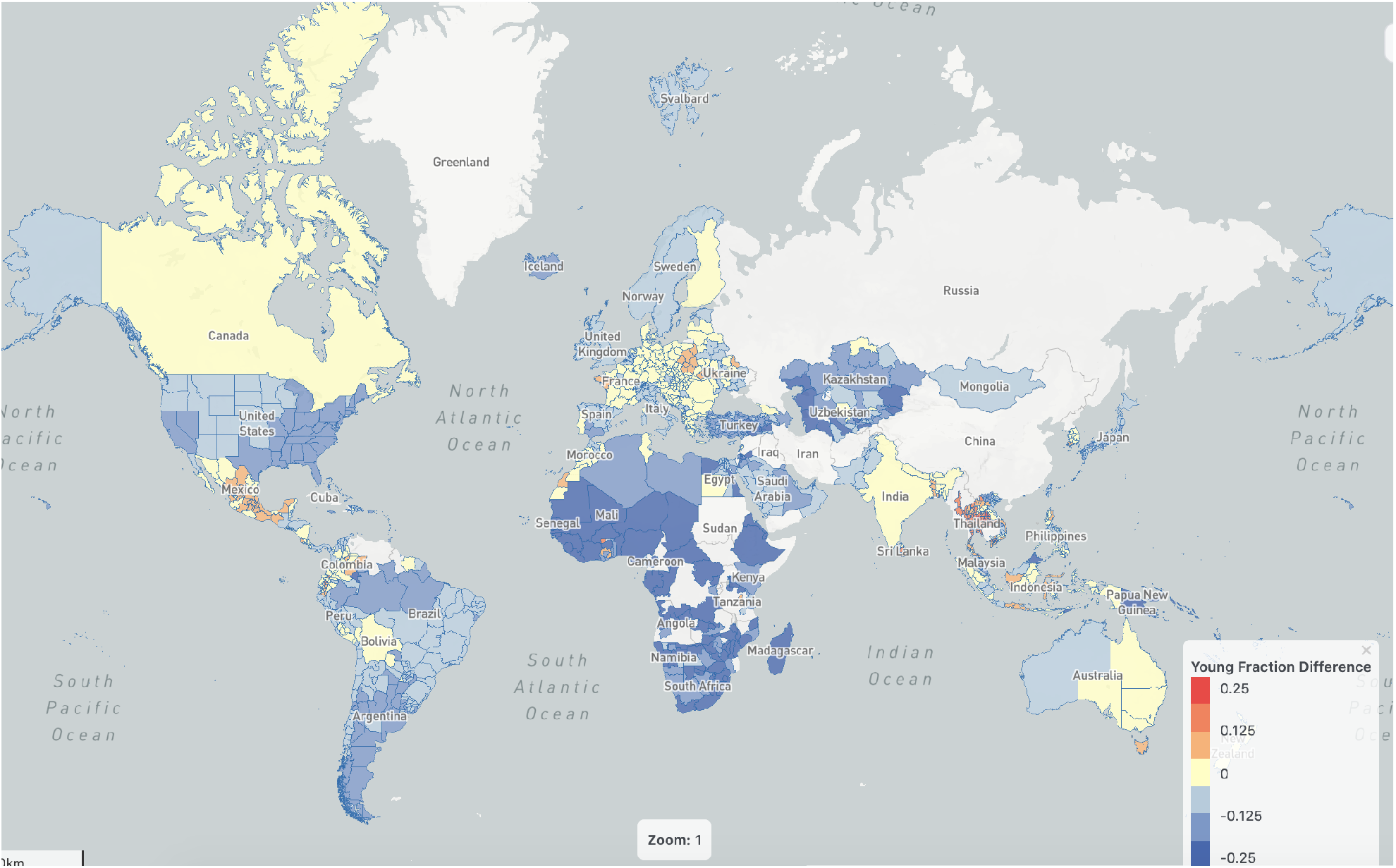
A global map that compares the age distribution of Colocation Map population and the overall population, July 2020. Red indicates that “young” fraction (defined as age between 15 and 29) is higher in Colocation Map population than in the HRSL and UN data, while blue indicates that “young” fraction is lower in Colocation Map population.

### 4.2 Within-Region Colocation vs. Between-Region Colocation

In a metapopulation model, the population associated with a given administrative polygon may interact very differently with itself than with other populations. Figure 5 shows that the colocation probabilities *p*_*rs*_ are indeed substantively different within-region than between-region. We observe that the scale of within-region colocation is orders of magnitude larger than between-region colocation. Next, there is not a clear behavior for between-region colocation versus the product of trajectory counts on the x-axis. The diagonal line of between-region points corresponds to samples where *m*_*rs*_ ≈ 1. On the other hand, the within-region points appear to scale as a function of the product of trajectory counts. The three lines, linear on the log-scale, correspond to the colocation probability scaling like 1*/n*, 1*/n*^3*/*4^, and 1*/n*^1*/*2^ respectively. If the colocation network was sparse within polygons (i.e. *m*_*rr*_ ∝ *n*_*r*_), we would expect a 1*/n* scaling. Instead we see that colocation within-regions appears more dense.

**Figure 5.**
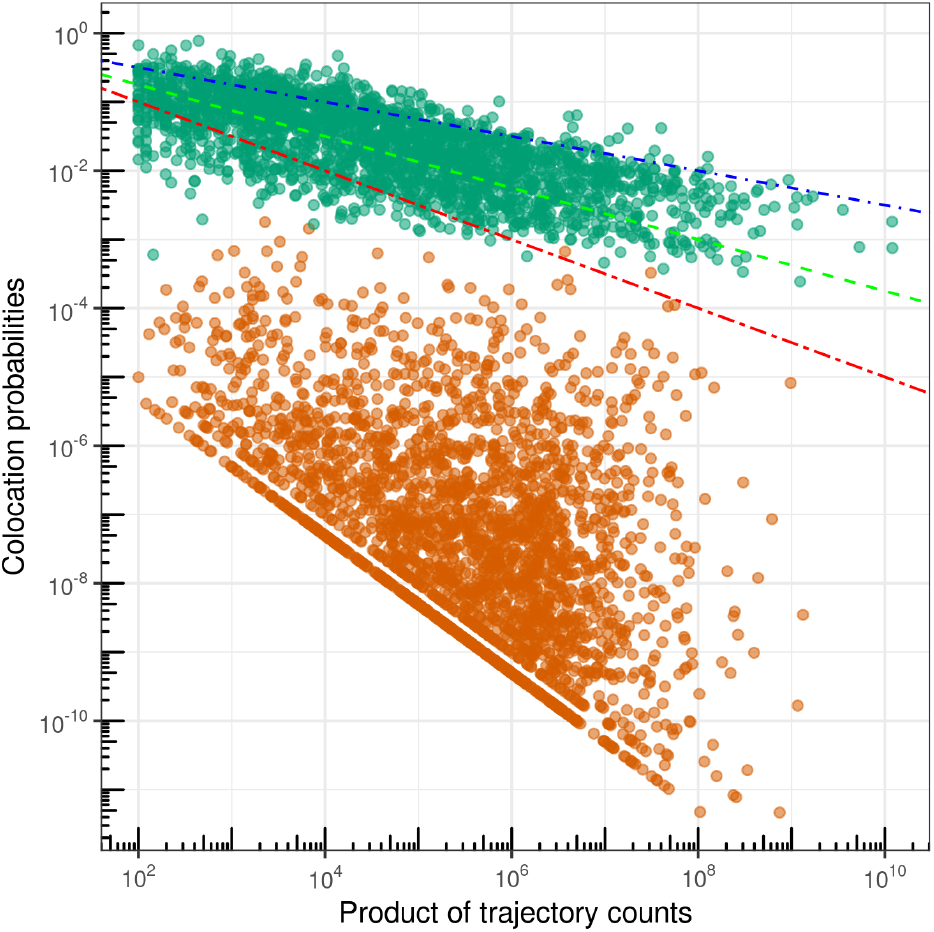
Samples (from 2020-04-07) of scaled within-(green) and between-polygon (orange) colocation probabilities *p*_*rs*_ on a log-scale versus the product of trajectory counts *n*_*r*_*n*_*s*_. Lines indicate a scaling of the within-region colocation probability as 1*/n* (two-dash red), 1*/n*^3*/*4^ (dashed green), and 1*/n*^1*/*2^ (dot-dash blue).

These observations may reflect the fact that within-region colocation can happen for qualitatively different reasons than between-region colocation. For example, two users that live in the same apartment building might always be colocated in the same Bing tile at night, and perhaps also during the day (especially during the COVID-19 pandemic, when many people are working from home). This points to a potential problem with Colocation Maps datasets, because people are unlikely to transmit infectious diseases between one another when they are simply sleeping in different rooms in an apartment complex. We will explore this issue further below.

### 4.3 Contact Heterogeneity

While Figure 5 explored how colocation rates vary over pairs of regions, we now examine heterogeneity in colocation over pairs of people. Figure 6 plots cumulative distributions of contact time over pairs. Consider any pair of people *i* and *j* who are ever colocated over the course of a week. To each such pair, we can assign the total number of minutes for which they are colocated, and we can also classify the pair as within-region (i.e., *i* and *j*’s homes are the same region) or between-region. The red and blue lines show the cumulative distributions of total colocation time over between-region and within-region pairs respectively. We find that the vast majority of pairs are colocated for very short periods of time, but there are a few that are colocated for many days over the course of a week.

**Figure 6.**
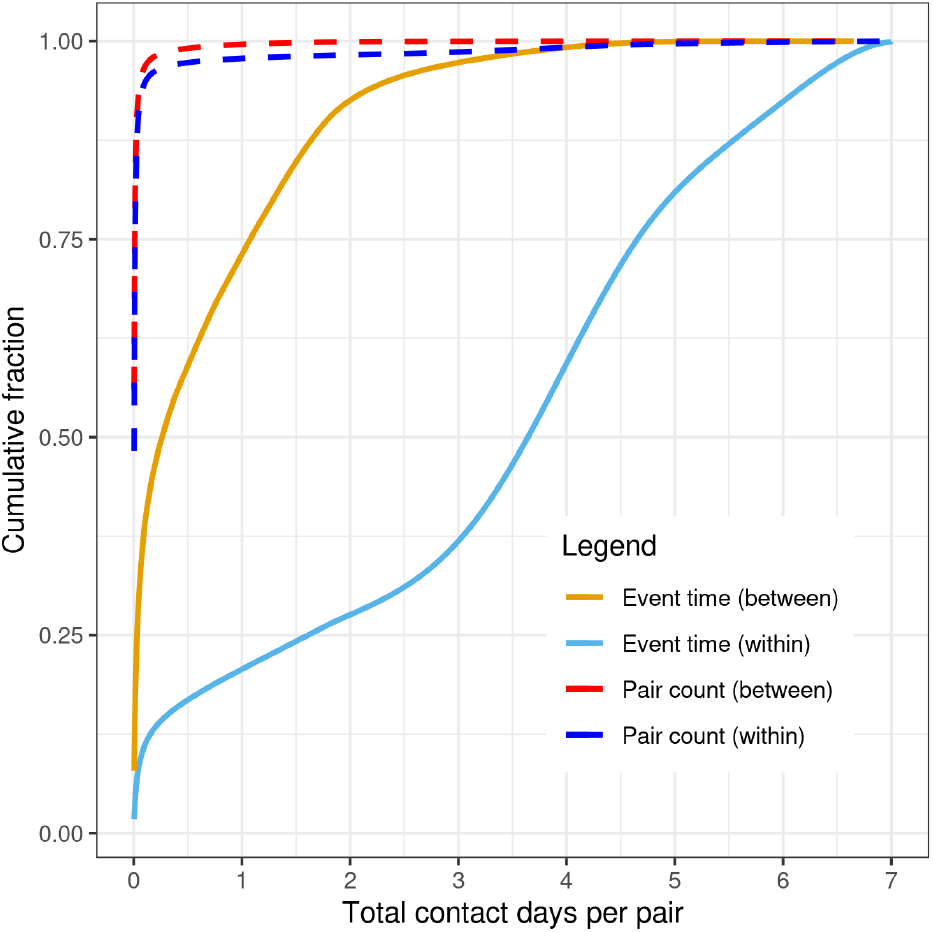
In dark blue and dark red respectively, the fractions of within-region pairs and between-region pairs accounted for by pairs that are colocated up to a certain amount of time. In teal and orange respectively, the fractions of overall within-region and between-region colocation time accounted for by pairs that are colocated up to a certain amount of time.

A pair that is colocated for many days contributes more to the overall colocation time (colocation minutes summed over all pairs) than a pair that is colocated for a very short period of time. To gain a better understanding of this, in Figure 6, we also plot the fraction of the total between-region (orange data points) and within-region (teal data points) colocation time that is accounted for by pairs that are colocated for up to a certain amount of time. We find that these distributions are broad, with the small percentage of pairs that are colocated for many days accounting for a substantial fraction of overall colocation time. In the case of between-region time, 75% of the overall colocation time comes from pairs that are colocated for less than a day, but that still leaves a quarter of the colocation time to come from pairs that are colocated for more than a full day over the course of the week. For within-region time, pairs that are colocated for long periods of time account for even more of the overall colocation time, with approximately a quarter of the time accounted for by pairs that are colocated for more than six days over the course of the week.

These observations point to considerable heterogeneity amongst pairs of people in the amount of time that they spend colocated. Network models or agent-based models would need to account for this heterogeneity to provide incremental value on top of metapopulation models. The differences between within-polygon and between-polygon behavior in Figure 6 also echo the potential problem with within-polygon colocation that we discussed above: within-polygon colocation time may be greatly affected by time that individuals spend sleeping or working in the same building, while not coming into contact that could result in infectious disease transmission.

### 4.4 Heterogeneity over Time

Colocation Maps aggregate colocation rates at the level of weeks. This is analogous to the situation with essentially any aggregate mobility dataset, where individual interactions are not reported alongside precise times, but it washes away ordering of contacts within the week. For example, it could be the case that colocation between two specific regions tends to happen earlier in the week and then fade away as the week progresses, with people from the two regions instead interacting more strongly with other regions. Such an interaction pattern could indeed affect the course of disease spread.

Do we see such interaction patterns in colocation data? Figure 7 shows how the colocation probabilities within and between the provinces of Torino and Milano in Italy vary hour-by-hour over the course of the week from Wednesday morning of July 8, 2020 through Wednesday morning of July 15, 2020. We see fluctuations over the course of each day, with colocation probabilities reaching a maximum within each province at night, when users are likely to be at home asleep. Within-region colocation probabilities reach a minimum during the day on weekdays. On Saturday and Sunday, however, we see slightly different behavior; most noticeably, the colocation between Torino and Milano increases on the weekends, perhaps reflecting weekend travel between the nearby regions. These properties of our data might affect epidemiological modeling efforts as well. We note how a large fraction of within-region colocation events appear to occur at night between 6 PM and 6 AM. It is reasonable to speculate that at this time many users are at home and interacting with members of their own households. Thus, the elevated within-region colocation scores do not necessarily reflect homogeneous mixing between individuals from different households, and it may be that including the nighttime colocation events exaggerate the average within-region colocation score.

**Figure 7.**
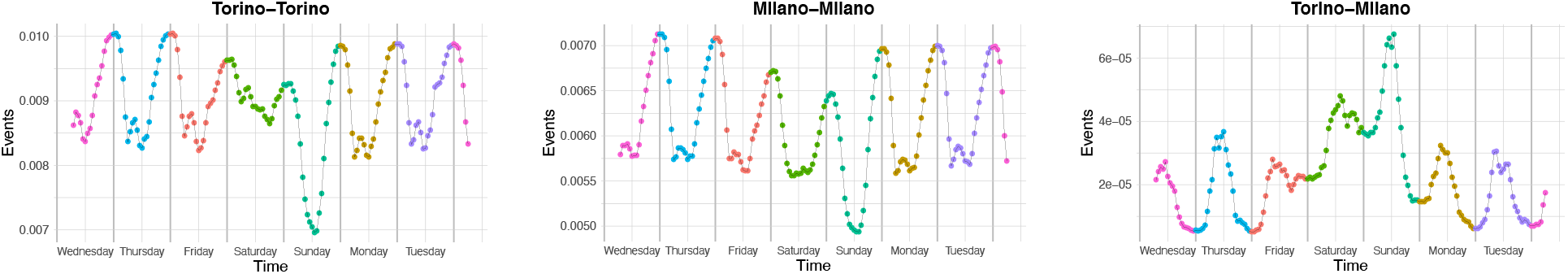
Colocation probabilities within and between the provinces of Torino and Milano in Italy, broken down by hour over the course of the week beginning Wednesday morning, July 8, 2020 and ending Wednesday morning, July 15, 2020.

### 4.5 Colocation vs. Face-to-Face Contact

A final issue is the distinction between disease-causing contacts and colocation. In order for a disease to be transmitted between individuals, very close proximity is often necessary, so that, for example, an uninfected individual could encounter a cough from an infected individual [52]. Because droplets from a cough do not travel half a kilometer, Colocation Maps are measuring colocation over a substantially larger spatial scale than would be needed to directly measure disease-causing contacts. Measuring physical proximity at the scale of a few meters would be technically difficult and, unless care is taken to obtain permissions from a willing population (as in [32]), possibly invasive. The perspective in Colocation Maps, then, is to get closer to the underlying network of possible disease-transmission events than is possible with traditional mobility data sources and to thereby enable better metapopulation modeling, while still respecting technical and ethical limitations.

Nevertheless, for end users of these datasets, it is important to understand the differences between “possible contacts” (which are aggregated into Colocation Maps) and “actual contacts” (to which we do not have access). Clearly, for a fixed set of individuals, the network of possible contacts will be much denser than the network of actual contacts. One interesting question, however, is the extent to which the edge weights in those two networks are correlated. We do not have the means to address this question with our data, but it has been addressed previously in the literature, by Génois and Barrat [32]. These authors compared face-to-face contact networks between people who consented to using wearable sensors in various contexts (schools, offices, hospitals, academic conferences) and colocation networks (measured at a 30 meter scale) for the same people. Génois and Barrat computed cosine similarities^9^ between the face-to-face contact and colocation networks and found reasonable values (∼0.7) across the different social contexts. These authors also found that individual-level properties (e.g., ordering of people based on contact degree) were *not* well conserved by the colocation networks. For our purposes, where we are not interested in sharing individual-level information and in fact want it to be as obscured as possible, this is an acceptable limitation. Génois and Barrat also found that downsampling approaches (where some colocation events are treated as face-to-face contacts and others are discarded) can help recover more realistic face-to-face contact networks and boost cosine similarity scores with the measured face-to-face contact network (up to the 0.9 range, depending on social context). We will discuss this possibility of downsampling our data in the Future Directions section below [32].

## Data Availability

The data used in this paper is made available through Facebook's Data for Good program under the terms of a Data License Agreement for research purposes. Any interested party can request access to this data by going to dataforgood.fb.com or by emailing.

https://dataforgood.fb.com

## 5. Future Directions

In this paper, we have described the construction of Facebook Colocation Maps and showed how they can facilitate improved metapopulation modeling of infectious diseases. Meanwhile, we have also critically examined the assumptions that underlie Colocation Maps, identifying certain discrepancies between the colocation rates in our datasets and the “real” networks of contacts that these datasets are meant to approximate. Although we have argued that our current Colocation Maps approximate the network of potential disease-transmission events better than usual mobility datasets (e.g., community flows), our explorations in the Assumptions of Colocation Maps section do motivate future improvements to these datasets. Here we will enumerate several directions for this work.

Assuming most people spend more time in their homes than any other locations, we should expect dramatically more colocation among people that live in the same tile. If those people share a household, then frequent contact is likely. But, when multiple homes or, especially, dense apartment buildings fall into the same tile, much of the observed colocation will be while people are sleeping and contagious disease transmission is unlikely. In the calculation of the final colocation rates, it may be worthwhile to subtract out colocation events that an individual logs within their own home region at night.

Additional signals about the context for a colocation event can be difficult to collect but could also improve the applicability of these datasets for modeling infectious-disease transmission. For example, whether colocation occurs indoors or outdoors can be relevant to the chances that the colocation leads to disease transmission. Satellite data, or other data on the fraction of a region that is covered by buildings, could be incorporated in the colocation calculation to weight colocation events based on whether they are likely to have occurred indoors or outdoors. Similarly, satellite data can help identify colocation events that happen on roadways, where people who are colocated because they are traveling in different vehicles are unlikely to transmit infectious diseases. Finally, social network data could be useful for better understanding colocations. For example, colocation events between people that know each other may be more likely to involve physical contact or close proximity than colocations between strangers, and the former could be upweighted in the calculation of the colocation rates.

Génois and Barrat have shown that downsampling colocation networks can yield contact networks that more closely approximate face-to-face contact networks [32]. This approach of downsampling the observed colocation events may provide an unifying framework for incorporating many of the considerations above. For example, it may be possible to retain each colocation event with a probability that depends upon:

- whether the colocation event happens at night between people from the same region
- whether the colocation event happens in a tile that contains a lot of indoor space
- whether the colocation event happens between people who are likely to know one another (e.g., Facebook friends)

This would amount to constructing a model that predicts when a colocation event is likely to correspond to close contact. This is a challenging task, but one that could improve the applicability of Colocation Maps to studies of infectious-disease transmission.

One limitation of our work is that biases in the underlying data, as well as those introduced via our trajectory filtering steps, have an unclear effect on colocation probabilities. The source data is a convenience sample of locations from Facebook users with smartphones who have opted into the Location History and background data collection features. Future work could provide a better understanding of how representative this population is, the implications of the biases on colocation probability, and an exploration of methods for bias correction.

## Acknowledgments

Thanks to Alessandro Vespignani, David Lazer, Adam Kucharski, and Oliver Brady for feedback on early versions of these datasets.

## Supplementary Material

### Derivation of SIR Dynamical Equations Using Colocation Probabilities

In the “Applications of Colocation Maps” section of the main paper, we provide a derivation of a metapopulation SIR model in terms of the parameters measured by Colocation Maps. Here, we provide some details of the derivation of the dynamical equations (6). These derivations use the assumption of homogeneity over individuals within subpopulations (see equation (6) in the main paper) and the assumption that colocation rates are constant over the week-long colocation period (i.e., that *m*_*rs,k*_ ≈ *p*_*rs*_*n*_*r*_*n*_*s*_):

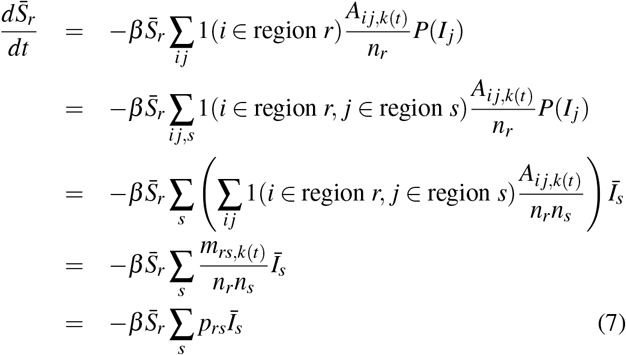

**Table S1.** Columns of the dataset that is shared through GeoInsights

| Column Name | Column Meaning |
| --- | --- |
| polygon1_id | a unique id for region 1 |
| polygon1_name | the name of region 1 |
| lon_1 | the longitude of the centroid of region 1 |
| lat_1 | the latitude of the centroid of region 1 |
| fb_population_1 | the number of people who were included in the calculation for region 1 |
| polygon2_id | a unique id for region 2 |
| polygon2_name | the name of region 2 |
| lon_2 | the longitude of the centroid of region 2 |
| lat_2 | the latitude of the centroid of region 2 |
| fb_population_2 | the number of people who were included in the calculation for region 2 |
| link_value | average fraction of the 1-week period in which pairs from regions 1 and 2 are colocated |

**Figure S1.**
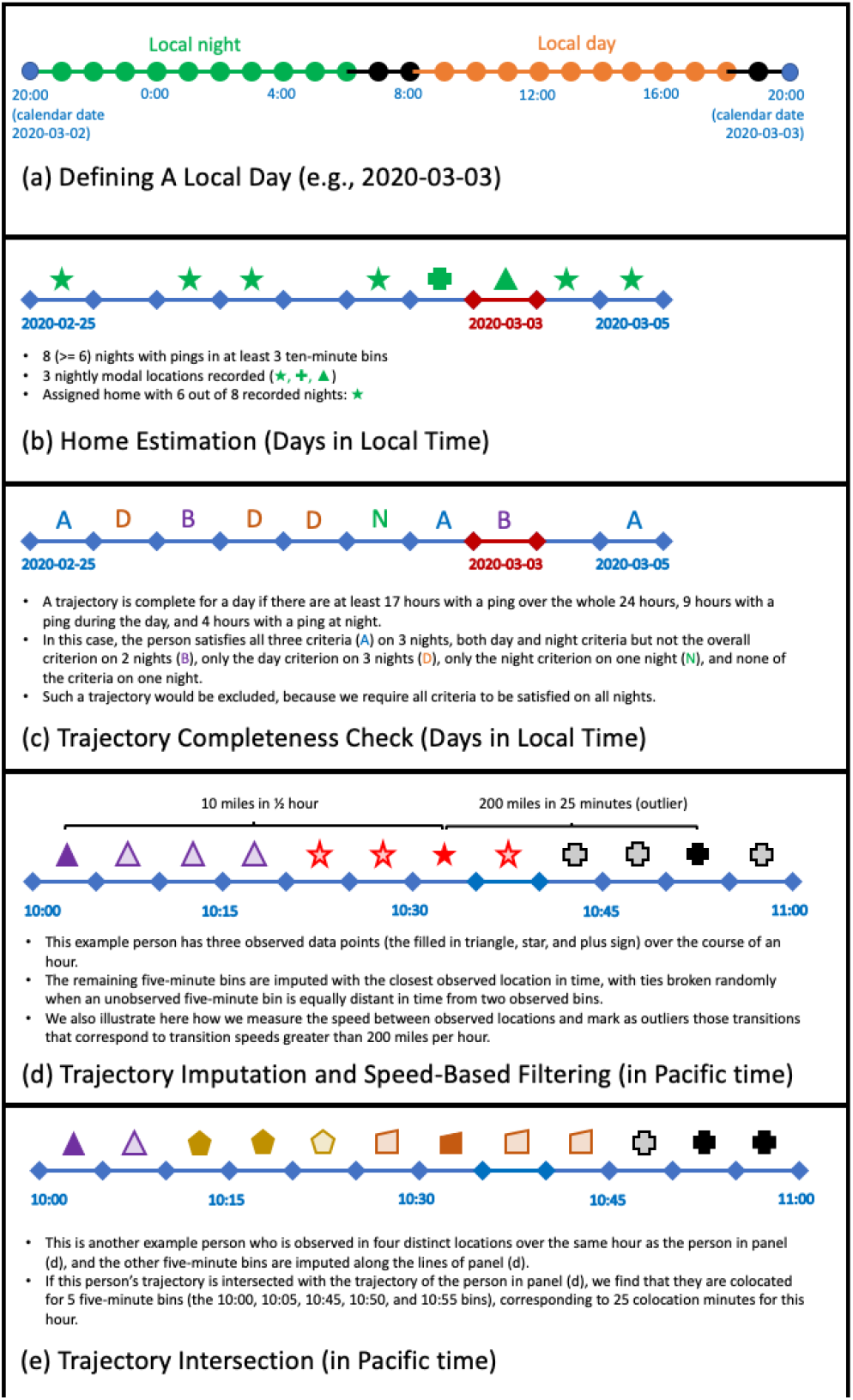
Details of calculation

**Figure S2.**
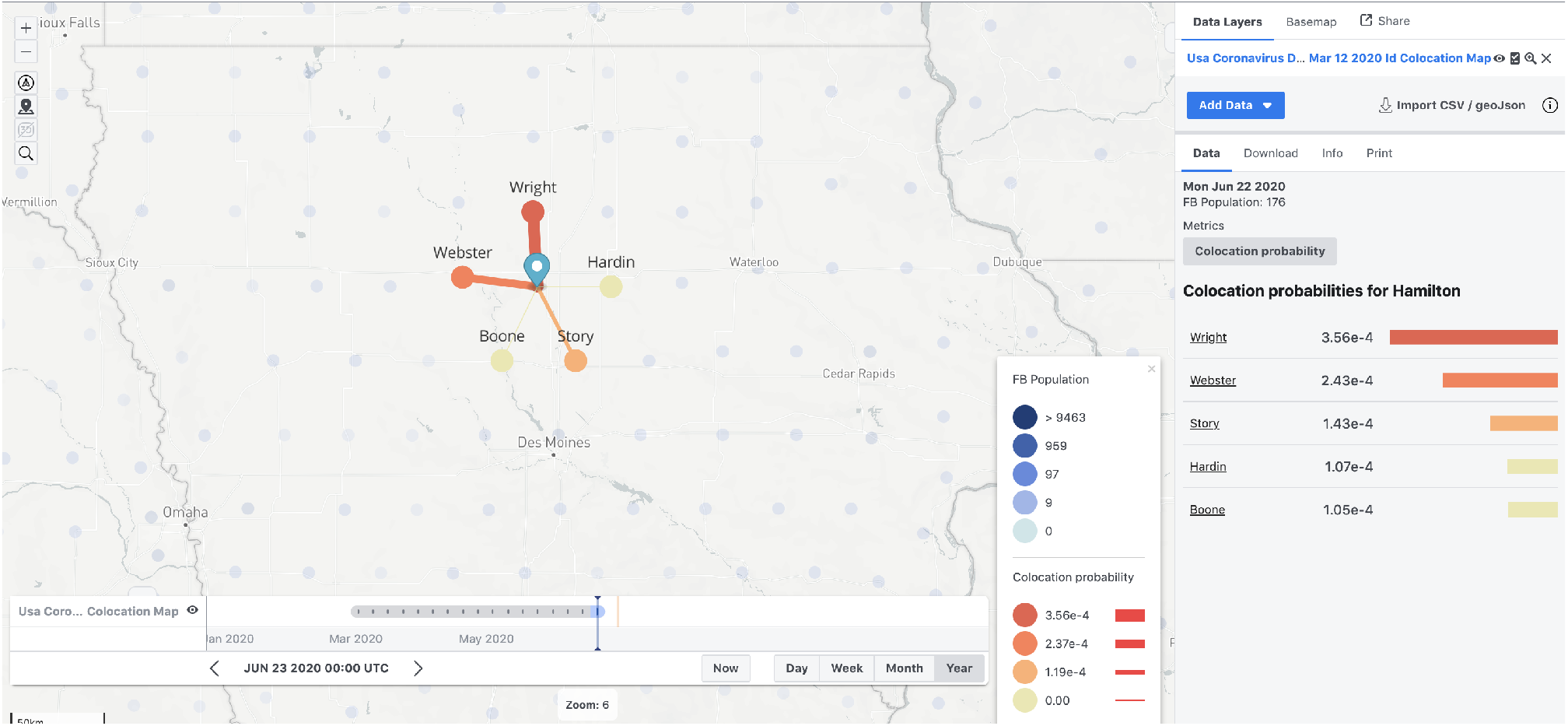
Example visualization of Colocation Maps in GeoInsights. This example shows strong connections for Hamilton County, Iowa in the period June 16 - June 22, 2020.

**Figure S3.**
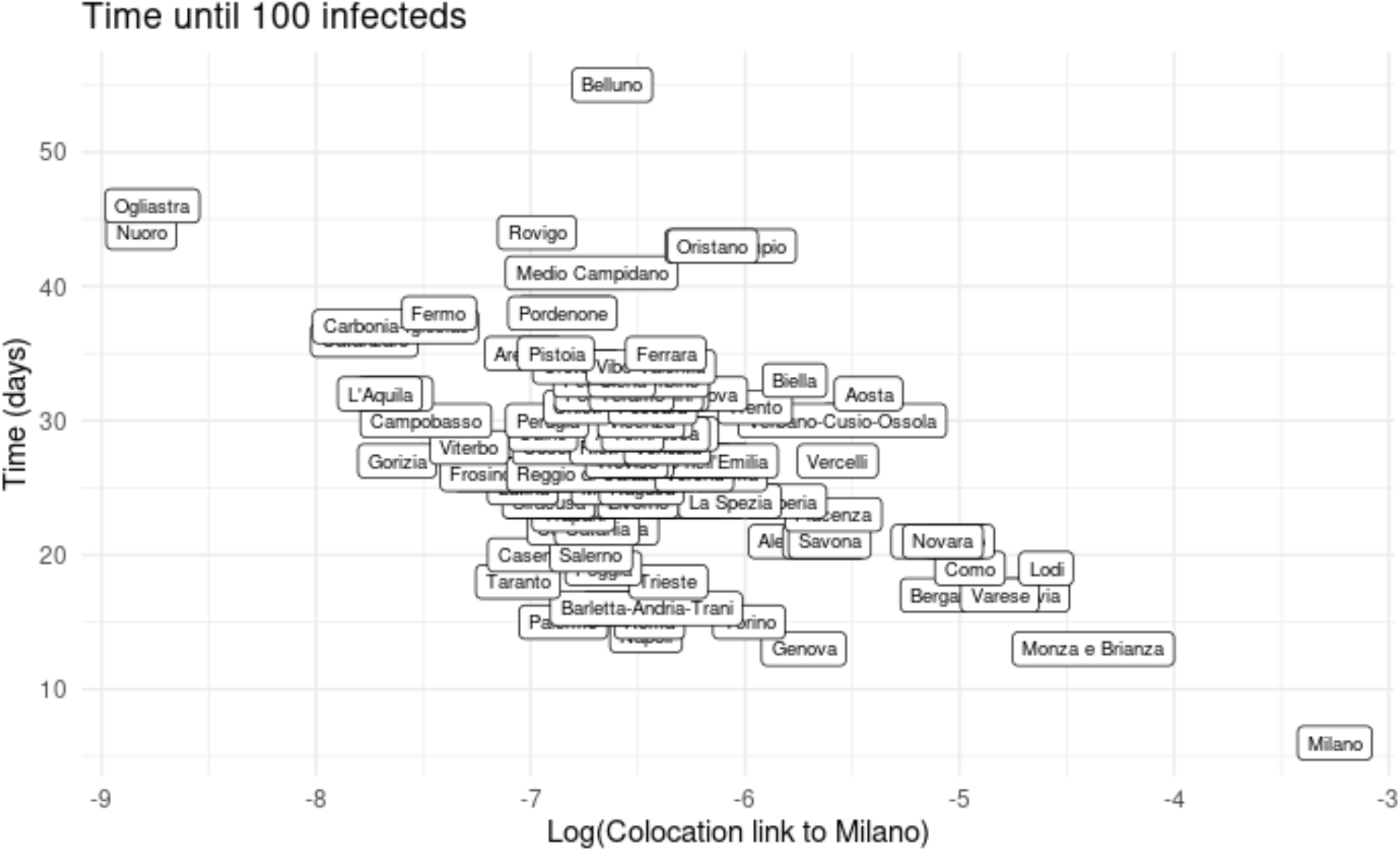
Time to 100 infections for the simulated outbreak shown in Figure 2 in the main text. The simulated outbreak starts in Milano. The x-axis shows the log of the colocation strength between each province and Milano where the outbreak starts. The y-axis shows the number of days until the number of infections reaches 100. The negative correlation suggests that the disease spreads more quickly to provinces which interact more strongly with the source of the outbreak.

**Figure S4.**
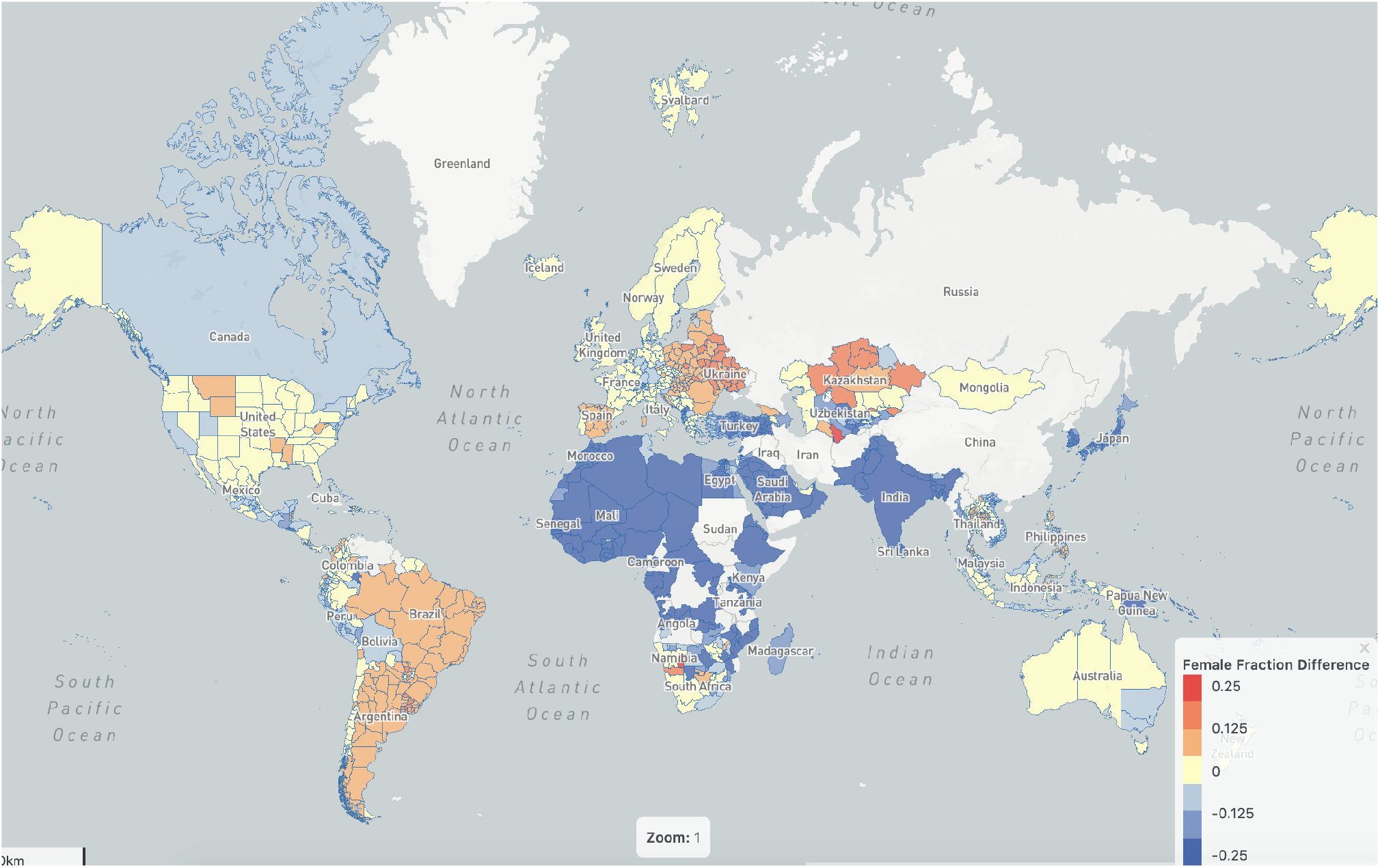
A global map that compares the gender distribution of Colocation Map population and the overall population, July 2020. Red indicates that female fraction is higher in Colocation Map population than in the HRSL and UN data, while blue indicates that female fraction is lower in Colocation Map population.

### Footnotes

1 For more details on the Location History setting, please see: https://www.facebook.com/help/278928889350358

2 Those interested in requesting access to Colocation Maps, or any other Facebook Data for Good maps, can do so via the instructions provided on the program’s website (https://dataforgood.fb.com/). The program website also maintains a collection of research publications which have made use of Colocation Maps and other resources to aid those seeking to make use of the data for their crisis-response work.

3 In practice, we recommend inserting another unknown parameter *κ* such that *m*_*rs,k*_ ≈ *n*_*r*_*n*_*s*_(1 + *δ*_*rs*_*κ*_*r*_)*p*_*rs*_ to rescale the diagonal values of *p*_*rs*_ (where the Kronecker delta *δ*_*rs*_ = 1 if *r* = *s* and zero otherwise). We suggest this modification because within-region colocation tends to occur for different reasons than between-region colocation. Within-region colocation is generally much larger than between-region colocation, and can be driven by nighttime contributions. This additional unknown parameter can help account for this difference in scale. See 4.2 and 4.4 for more details.

4 http://maths.adelaide.edu.au/lewis.mitchell/socialdistancing Last time visited on July 27th 2020.

5 https://chapman.maps.arcgis.com/apps/opsdashboard/index.html/d5d4047f03b742e9bddb2af75e5b9ba8 Last time visited on July 27th 2020.

6 https://chapman.maps.arcgis.com/apps/opsdashboard/index.html/3eb4582a297d42d498066309668427bf Last time visited on July 27th 2020.

7 https://cmmid.github.io/colocation_dashboard_cmmid/ Last time visited on July 27th 2020.

8 https://population.un.org/wpp/DataQuery/ Last time visited on August 6th 2020.

9 Génois and Barrat compute cosine similarities between adjacency matrices by simply concatenating the rows of the matrices into vectors [32].

